# Characterizing proteomic and transcriptomic features of missense variants in amyotrophic lateral sclerosis genes

**DOI:** 10.1101/2022.12.21.22283728

**Authors:** Allison A. Dilliott, Guy A. Rouleau, Sumaiya Iqbal, Sali M.K. Farhan

## Abstract

**Background:** Within recent years, there has been a growing number of genes associated with amyotrophic lateral sclerosis (ALS), resulting in an increasing number of novel variants, particularly missense variants, many of which are of unknown clinical significance. Here, we leverage the sequencing efforts of the ALS Knowledge Portal (3,864 individuals with ALS and 7,839 controls) and Project MinE ALS Sequencing Consortium (4,366 individuals with ALS and 1,832 controls) to perform proteomic and transcriptomic characterization of missense variants in 24 ALS-associated genes.

**Results:** Using predicted human protein structures from AlphaFold, we determined that missense variants carried by individuals with ALS were significantly enriched in β-sheets and α-helices, as well as in core, buried, or moderately buried regions. At the same time, we identified that hydrophobic amino acid residues, compositionally biased protein regions and protein-protein interaction regions are predominantly enriched in missense variants carried by individuals with ALS. Assessment of expression level based on transcriptomics also revealed enrichment of variants of high and medium expression across all tissues and within the brain. We further explored enriched features of interest using burden analyses to determine whether individual genes were driving the enrichment signal. A case study is presented for *SOD1* to demonstrate proof of concept of how enriched features may aid in defining variant pathogenicity.

**Conclusions:** Our results present proteomic and transcriptomic features that are important indicators of missense variant pathogenicity in ALS and are distinct from features associated with neurodevelopmental disorders.

## Background

Amyotrophic lateral sclerosis (ALS), also referred to as motor neuron disease, is a fatal neurodegenerative disorder characterized by adult-onset progressive degeneration of upper and lower motor neurons (1). The neuronal loss leads to progressive muscle weakness and eventual paralysis and death within three-five years predominantly due to respiratory failure. Importantly, ALS displays high heritability, not only for individuals with familial forms of the disease, but also for seemingly sporadic or isolated cases with no apparent family history (2). The most common genetic causes of ALS, namely pathogenic variants in *C9orf72, SOD1, TARDBP*, and *FUS*, have been reported in both patients with familial ALS (fALS) and sporadic ALS (sALS) (3-5). As such, genetic testing has become an important tool in the clinical care of ALS patients, offering many advantages, including early, accurate diagnosis and access to emerging clinical trials targeted to specific genetic profiles (6-8). These benefits are now considered quite substantial and recent recommendations include the expansion of clinical genetic testing to a broader range of patients, not only to those with a known family history as has been the conventional approach (9).

However, standardized clinical testing in ALS remains challenging. Over recent years, there has been a concerted effort to uncover novel genetic markers associated with ALS with a growing number of genes discovered (10), yet there are a large proportion of patients that remain genetically unexplained (5). As the compendium of ALS associated genes has expanded, there has been a vast increase in the number of novel variants identified in those genes, and consequently, accurate pathogenicity classification remains challenging. Typically, variants are classified based on the American College of Medical Genetics and Genomics (ACMG) pathogenicity classification guidelines, which has become a standardized approach applied to all diseases and take into consideration a variant’s previous clinical reports, functional experimental evidence, segregation within a pedigree, minor allele frequency in healthy populations and relevant disease cohorts, and *in silico* predictions, among other evidence (11). But in most cases, the sum of the evidence is not sufficient to confidently classify a variant as pathogenic or benign, and the classification of variants of uncertain significance (VUS) in clinical testing is common. Previous genetic testing of known genes in ALS cohorts have identified VUS in 15-25% of patients (12-15), and data obtained from the variant database, ClinVar, demonstrates that even the most common, well studied ALS associated genes — *SOD1, TARDBP*, and *FUS* — have relatively high rates of VUS (Figure 1A). Not only is the identification of VUS from clinical testing frustrating for the patient and their families, but it can complicate genetic counselling and prevent patient enrollment in ongoing clinical trials (16, 17).

**Figure 1.**
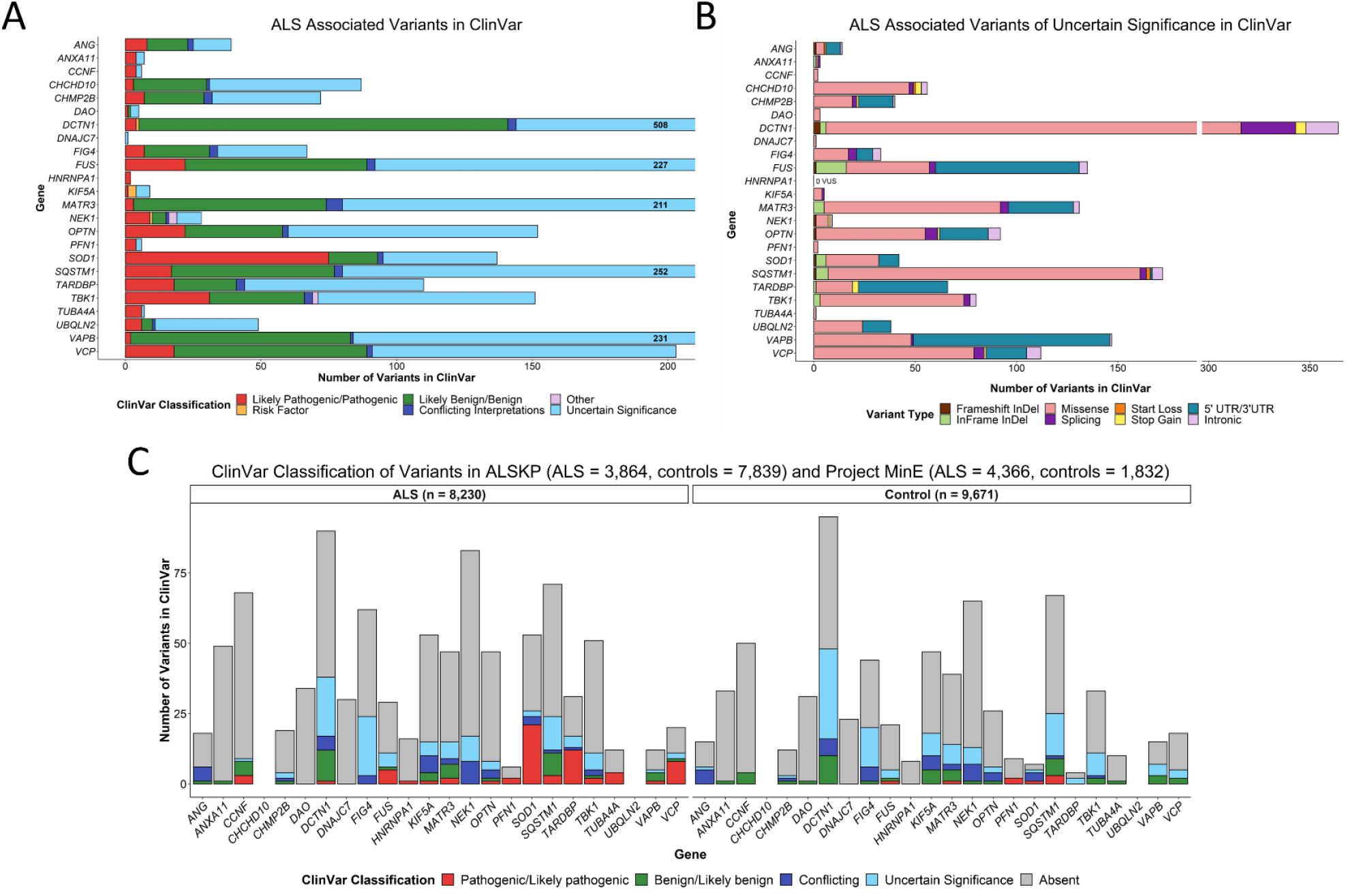
Pathogenicity classification and variant types reported in ClinVar in genes associated with ALS. **(A)** Pathogenicity classification of all variants reported in ClinVar as associated with ALS and/or motor neuron disease. The total number of variants reported in ClinVar in *DCTN1, FUS, MATR3, SQSTM1*, and *VAPB* exceeded the y-axis limit of 200; in these cases, the total number of variants found in ClinVar were included on the right side of the bar plot. **(B)** Variant types of all variants reported in ClinVar as being of uncertain significance for ALS and/or motor neuron disease. **(C)** Pathogenicity classification reported in ClinVar of variants identified across 24 ALS associated genes in the ALS Knowledge Portal (ALSKP) and Project MinE ALS sequencing consortium datasets.

Missense variants are particularly common VUS (18), as the variant consequence can be difficult to predict without definitive experimental evidence to determine whether function of the encoded protein is gained, lost, or altered. Importantly, missense variants are established drivers of neurodegenerative disease risk, as they can induce a gain of protein activity as observed in *PSEN1* and *PSEN2*, which cause early onset Alzheimer’s disease and *SOD1* in ALS (19-24). Although many *in silico* tools have been developed to aid in prediction of missense variant pathogenicity (25-28), the tools primarily rely on variant conservation and constraint, as well as broad biochemical properties. *In silico* tools are also not specific to any singular disease, such as ALS, and may fail to capture phenotype- and gene-specific nuances driving missense variant pathogenicity. Therefore, the tools and the ACMG guidelines may fail to capture evidence that could be derived from protein structure and function or from transcriptional expression.

Recently, enrichment analyses were applied to identify protein structural features characterizing likely pathogenic variation in neurodevelopmental disorders (NDD) (29). Here, we aim to expand upon this approach to characterize the amino acid residue changes resulting from genetic missense variation in known ALS associated genes, observed in large cohorts of ALS patients and controls, to identify signals of enrichment of specific protein structure, protein sequence, and transcriptomic features. We anticipate that our results will contribute to the improvement of interpretation of VUS pathogenicity in ALS clinical testing.

## Methods

### ALS associated gene selection

In total, 24 genes were selected that had been previously associated with ALS in the literature. Specifically, we only included genes with at least two publications describing rare coding single nucleotide variants in individuals with ALS. We also excluded any genes that display an unclear inheritance pattern in ALS (i.e. driven by risk alleles from genome-wide association studies) or had a limited or refuted gene-disease validity classification from the ALS ClinGen Gene Curation Expert Panel as of January 2022 (Clinical Genome ALS). Summary of the 24 ALS-associated genes, including the year of first association, inheritance pattern, method of first association, and protein function, is presented in Supplemental Table 1.

### Study samples and sequencing

Sequencing data were obtained from the ALS Knowledge Portal and Project MinE ALS sequencing consortium (30, 31). Briefly, the ALS Knowledge Portal contains summary data from whole exomes of 3,864 individuals with ALS and 7,839 controls. The dataset was subjected to rigorous and standard quality control, including variant and sample level assessment to account for depth, coverage, genetic ancestry matching using principal component analysis (PCA), and relatedness using identity by descent metrics. Carriers of the *C9orf72* hexanucleotide expansion were excluded from the study. Further, approximately 2,000 of the case samples were screened for pathogenic variants in *SOD1, FUS*, and *TARDBP*, and known variant carriers were excluded. Comprehensive details outlining sample acquisition, exome-sequencing, and quality control were previously described (30). The Project MinE ALS sequencing consortium dataset contains whole-genome sequencing of 4,366 individuals with ALS and 1,832 controls. Again, details regarding sequencing methodology and quality control were previously described and largely compare to the steps undertaken by the ALS Knowledge Portal (31). Controls were matched to the individuals with ALS based on age, sex, and geographical region. For the purposes of this study, Project MinE data were obtained through the Project MinE Data Browser^1^.

### Variant genomic annotation and filtering

Variants from both datasets were annotated using Variant Effect Predictor (Ensemble v.105.0) (32) with the following features: gene symbol; Human Genome Variation Society (HGVS) coding and protein sequence alteration; variant consequence; Genome Aggregation Database (gnomAD) v2.1.1 Non-Finnish European (NFE), non-neurological (n = 51,592) allele frequency (33); and ClinVar pathogenicity classifications (34). Only variants within the 24 ALS-associated genes were retained for further analysis. Variants were also filtered to only include missense variants that were rare in the general population (allele frequency < 0.01 in the gnomAD v2.2.2 NFE non-neurological cohort).

### Variant protein feature annotation

Amino acid residues substituted in missense variants were annotated by two sets of protein sequence features and two sets of protein three-dimensional (3D) structural features.

The protein sequence features included functional site annotations from the UniProt database (35) and post-translational modification (PTM) site annotations from the PhosphoSitePlus database (36), referred to as functional and PTM features, respectively. Affected amino acid residues in the identified missense variants were annotated with twenty-six functional features including active site, binding site, calcium binding site, coiled coil, compositional bias, cross link, disulphide bond, DNA binding site, domain, glycosylation, intramembrane, lipidation, metal binding site, modified residue, motif, nucleoprotein binding site, peptide, propeptide, region of interest, repeat, signal peptide, site, topological domain, transit peptide, transmembrane, and zinc finger (Supplemental Table 2)^2^. Similarly, affected amino acid residues in the missense variants were annotated with ten PTM features including acetylation, glycosylation (O-glycosylation), methylation, phosphorylation, sumoylation, ubiquitination, regulatory site, kinase-substrate, PTMvar (i.e., PTMs overlapping genetic variants), and disease-associated PTM (Supplemental Table 3).

Predicted monomeric 3D structures of the 24 ALS associated proteins were collected from the AlphaFold database (37, 38) and were used to extract structural features. From the 3D structures, per-residue secondary structure type and residues’ solvent accessible surface area (ASA) were derived using the dictionary of protein secondary structure (DSSP) program (39). Each amino acid residue was annotated with one of nine different types of secondary structure. These nine different types of secondary structures were grouped into three broader categories: (i) β-strand/-sheet included β-strands and β-sheets; (ii) helices included 3_10_-helices, α-helices, π-helices, and polyproline II helices; and (iii) coils included loops, bends, and turns. For each residue, the ASA value (measured in units of square of Angstroms, Å^2^) was transformed to calculate the Relative ASA (RSA) by normalizing the ASA of that residue by the surface area of the same type of residue in a reference state. We used the ASA normalizing values derived in Tien et al. (40) using Gly-X-Gly tripeptide as the reference state for a given residue X. Based on the value of RSA, we labeled each amino acid with one of the following exposure levels: core, RSA < 5%; buried, 5% < RSA < 25%; medium-buried, 25% < RSA < 50%; medium-exposed, 50% < RSA < 75%; and exposed, RSA > 75%, as described by Iqbal et al. (41). Note that for the structural feature analyses involving the annotations obtained from AlphaFold predicted structures, only variants at residues with a high quality (predicted local-distance difference test [pLDDT] > 70; per-residue estimates of reliability generated by the AlphaFold neural (37)) were used.

### Transcriptional variant expression annotation

Finally, the missense variants were annotated based on the expression level using data obtained from the Genotype-Tissue Expression (GTEx) portal (42). Average expression of each amino acid across all GTEx tissues were calculated and hereafter referred to as “all tissues” expression levels, and average expression of each amino acid across all GTEx brain tissues were calculated and hereafter referred to as “brain” expression levels. Expression levels were binned based on the following: low, mean expression < 0.1; medium, mean expression ≥ 0.1 and ≤ 0.9; and high, mean expression > 0.9 as described by Cummings et al. (43).

### Statistical analyses

Missense variants were binned based on the above proteomic and transcriptomic features. To quantify the enrichment of missense variants with respect to these bins in the ALS Knowledge Portal and Project MinE ALS datasets, and identify features associated with ALS, we applied two-sided Fisher’s exact tests. Specifically, the number of individuals with ALS carrying each respective variant type was compared to the number of controls carrying the variant type. We also applied a combined analysis between the two cohorts using the Cochran-Mantel-Haenszel (CMH) test. Significance was measured at an alpha-level of 0.05.

For proteomic and transcriptomic features demonstrating significant enrichment in individuals with ALS compared to the controls, variants were further binned by genes and the Fisher’s exact test was again applied, to determine if specific genes were driving the enrichment signals. Similarly, individual tests were applied to the ALS Knowledge Portal and Project MinE ALS datasets, followed by combined analyses between the two cohorts using the CMH test. Following Bonferroni multiple testing correction to account for the 24 genes, significance was measured at an alpha level of 2.08e-03.

Statistical analyses were performed using the R statistical software v4.1.1 (44) in R Studio v1.4.1717. Data visualization was performed using the *ggplot2* R package (v3.3.6) (45).

### SOD1 case study

The Genomics 2 Proteins (G2P) portal^3^ was used to map *SOD1* missense variants, collected from the ALS Knowledge Portal and Project MinE sequencing consortium and classified according to ClinVar pathogenicity ascertainment, onto the SOD1AlphaFold predicted protein structure (AlphaFold ID: AF-P00441-F1) (41). Similarly, the protein sequence and structural features as well as transcriptomic features for each residue were mapped onto the structure of SOD1 using the G2P portal. The structure annotated with variants and features was exported from the portal and visualized using the PyMOL software for downstream analyses (i.e., investigating the overlap between residues mutated in pathogenic and VUS in individuals with ALS and residues with features significantly associated with ALS variants in *SOD1*).

## Results

### Missense variants identified in ALS associated genes

We first assessed the contribution of missense variation to the amount of uncertainty in pathogenicity assessment of ALS associated variation by interrogating the 24 selected ALS associated genes in the ClinVar database and identified all ALS associated variants within the genes (Figure 1A). *DCTN1* had the greatest number of VUS reported in ClinVar (n = 364), although five other genes each had >100 VUS reported, including *FUS, MATR3, SQSTM1, VAPB*, and *VCP*. In four out of these six genes with >100 VUS, the largest number of reported VUS in ClinVar were missense variants (Figure 1B).

Across 3,864 individuals with ALS and 7,839 controls from the ALS Knowledge Portal, 728 unique rare, missense variants were identified within 23 of the 24 ALS associated genes (Supplemental Table 4). No unique, rare variants were detected in *CHCHD10*. In 4,366 individuals with ALS and 1,832 controls from the Project MinE ALS sequencing consortium, 609 unique rare, missense variants were identified in 23 of the 24 ALS associated genes (Supplemental Table 4). However, in this dataset no variants were reported in *UBQLN2*.

Again, the ClinVar database was interrogated, specifically for the rare variants identified in the ALS Knowledge Portal and Project MinE ALS sequencing consortium datasets (Figure 1C). Similarly, the largest number of VUS in both the individuals with ALS and controls within our dataset were observed in *DCTN1*.

### Variant reference and alternate amino acids

We assessed the distribution of the reference amino acids involved in the rare missense variants identified in the individuals with ALS and controls of the ALS Knowledge Portal and Project MinE ALS sequencing consortium (Figure 2A). To determine which reference amino acids were more frequently impacted in ALS, an enrichment analysis was performed on both datasets, followed by a combined analysis including variants observed in both datasets. From the combined analysis, we observed a significant enrichment of rare, missense variants affecting arginine (Arg), asparagine (Asn), aspartic acid (Asp), glutamine (Gln), glycine (Gly), isoleucine (Ile), leucine (Leu), lysine (Lys), proline (Pro), and valine (Val) (p < 0.05; Figure 2B). We also assessed whether specific genes were driving these enrichments of rare, missense variants affecting specific amino acids and observed individuals with ALS to be significantly enriched for variants affecting Ile and Leu in *SOD1* (OR = 71.94 [4.40-1176.74], p = 1.19e-11 and OR = 20.00 [1.15-346.50], p = 1.85e-03, respectively); and for variants affecting Arg in both *NEK1* (OR = 1.85 [1.36-2.50], p = 9.15e-05) and *VCP* (OR = 20.99 [2.53-174.35], p = 3.01e-04) across the combined datasets (Supplemental Figure 1).

**Figure 2.**
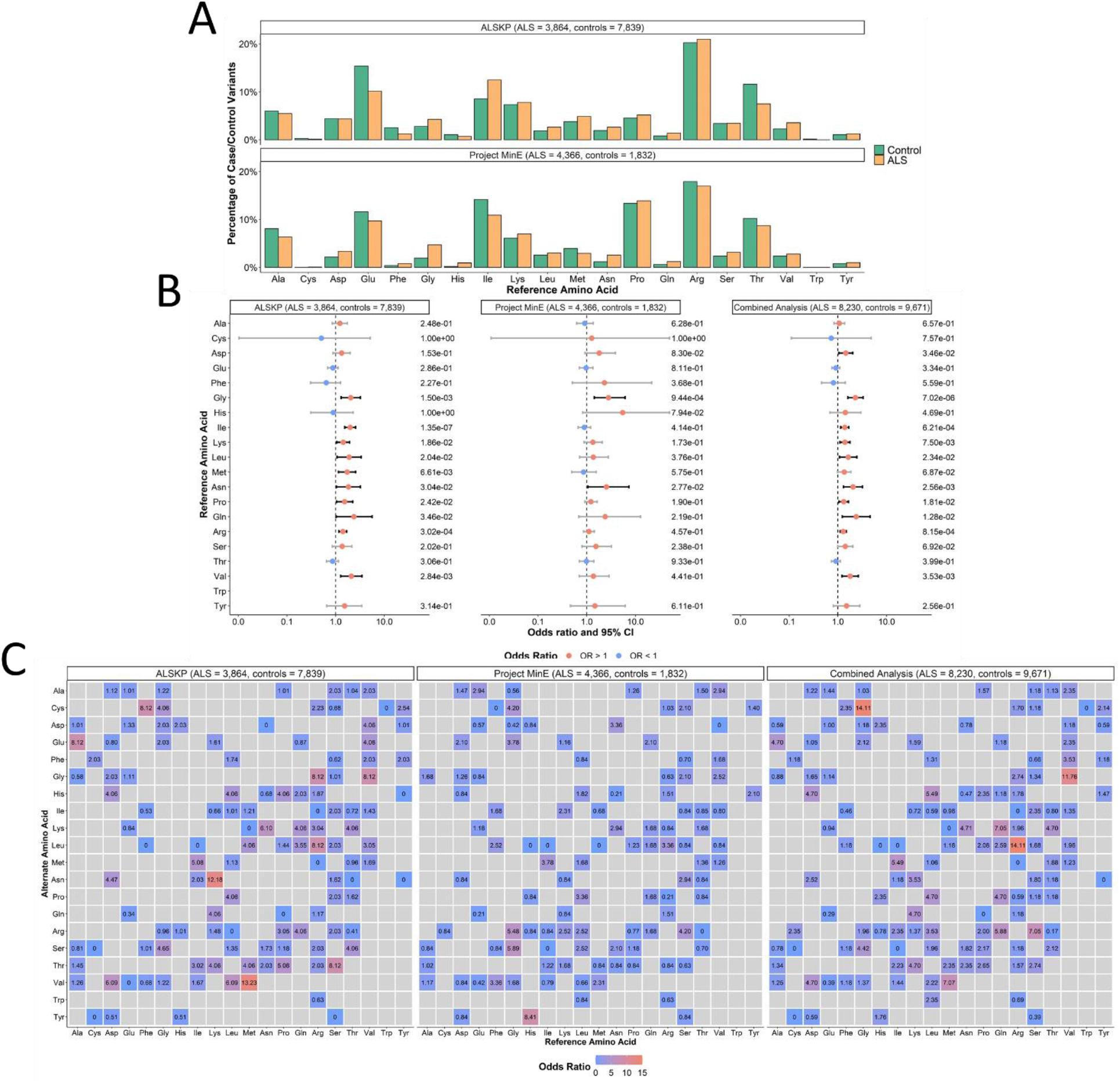
Distribution of amino acids involved in rare missense variants identified across 24 ALS associated genes. **(A)** Distribution of the amino acids involved in all variants of interest were identified in the ALS Knowledge Portal (ALSKP) and Project MinE ALS sequencing consortium datasets. **(B)** Enrichment analyses were performed using Fishers Exact testing to compare the number of variants carried by individuals with ALS and controls of affecting different amino acids in the ALSKP and Project MinE datasets, followed by a Cochran–Mantel– Haenszel (CMH) test on the combined datasets. Significance was measured at an alpha-level of 0.05. **(C)** Heat map displaying the reference and alternate amino acids of rare missense variants identified across 24 ALS associated genes in ALS case-control sequencing datasets, defined by the reference amino acid. Odds ratios were calculated to compare the frequency of the change in the individuals with ALS to the controls from the ALS Knowledge Portal, Project MinE ALS, and combined datasets.

We further examined the combinations of reference amino acids and alternate amino acids involved in the rare, missense variants in individuals with ALS versus controls. Across the combined datasets, the largest odds ratios were observed for missense variants involving Gly to cystine (Cys), Arg to Leu, or Val to Gly amino acid changes (OR = 14.11, 14.11, and 11.76, respectively; Figure 2C).

### Variant protein structural feature assessment

Quality of the predicted monomeric structures of the 24 ALS associated proteins collected from the AlphaFold database are presented in Supplemental Table 5. For analyses involving the annotations obtained from AlphaFold predicted structures, only variants at residues with a high quality (pLDDT > 70 (37)) were retained, including 74.2% of variants carried by individuals with ALS and 75.0% of variants carried by controls from the ALS Knowledge Portal; and 62.6% of variants carried by individuals with ALS and 64.5% of variants carried by controls from the Project MinE ALS sequencing consortium (Supplemental Figure 2).

From the AlphaFold annotations, we assessed the secondary structure distribution of the reference amino acids involved in the rare, missense variants (Figure 3A) to determine whether specific secondary structure types were more often affected in variants carried by individuals with ALS than controls (Figure 3B). Following binning of the secondary structures into sub-types, the combined dataset analysis revealed that variants at residues involved in β-strands/sheets (OR = 1.91 [1.51-2.42], p = 5.21e-08) and involved in helices (OR = 1.35 [1.21-1.50], p = 2.65e-08) were significantly enriched in ALS. Notably, these sub-type signals may have been driven by the significantly enriched secondary structure type signals identified for β-sheets (OR = 1.95 [1.53-2.47], p = 3.69e-08) and α-helices (OR = 1.32 [1.18-1.48], p = 3.84e-07; Supplemental Figure 3). The β-strands/sheets signals were primarily driven by *SOD1* (OR = 20.78 [7.14-60.47], p = 6.97e-14) and *ANG* (OR = 3.57 [1.65-7.72], p = 1.05e-03), whereas the signals in helices were driven by *NEK1* (OR = 1.80 [1.36-2.38], p = 4.81e-05), *SQSTM1* (OR = 1.57 [1.21-2.05], p = 6.41e-04), and *OPTN* (OR = 2.76 [1.49-5.09], p = 9.52e-04).

**Figure 3.**
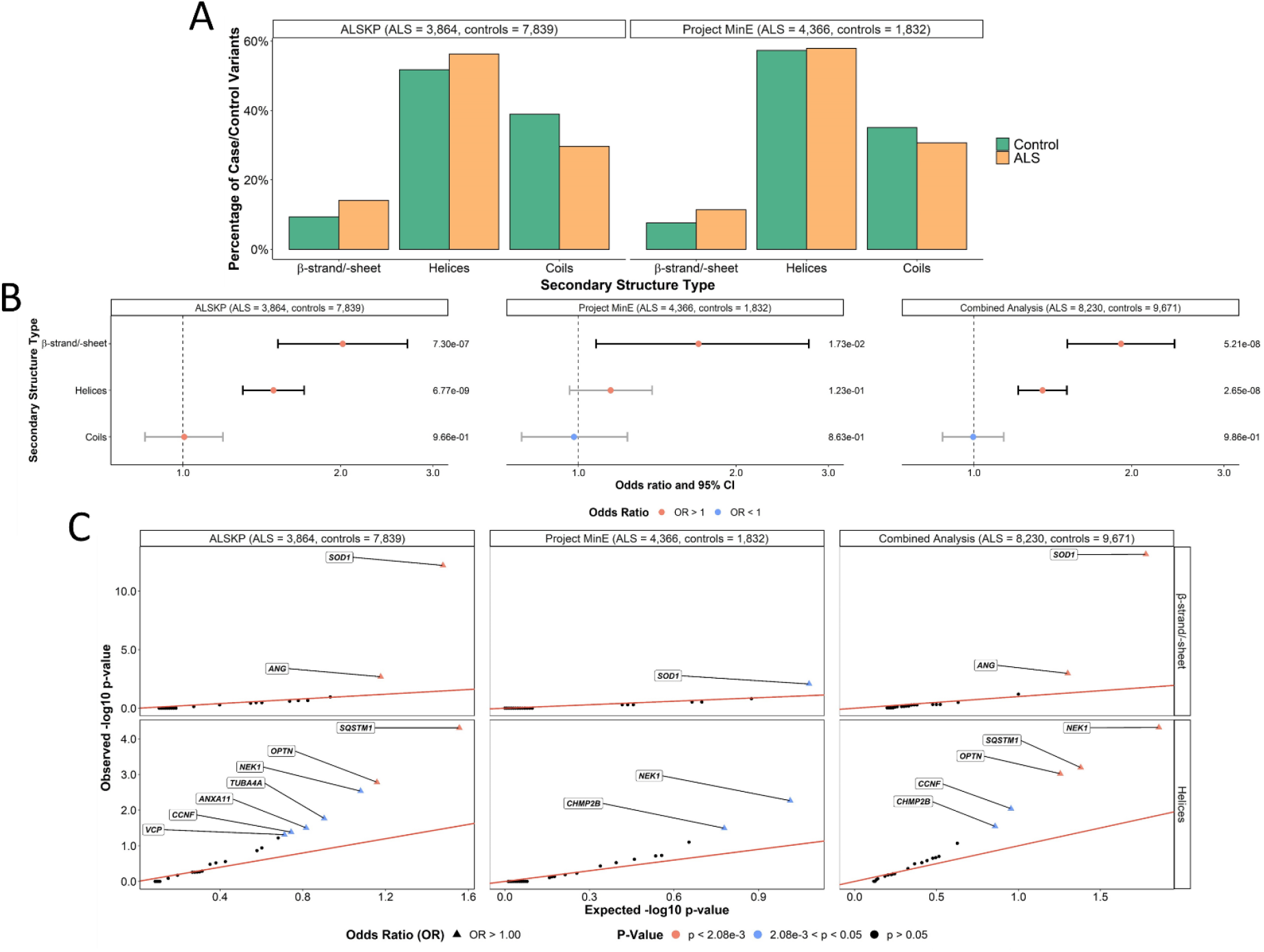
Secondary structure types at which rare missense variants were identified across 24 ALS associated genes. **(A)** Secondary structure types were obtained using the AlphaFold predicted structures and DSSP (dictionary of protein secondary structure) program for all residues at which variants of interest were observed in the ALS Knowledge Portal (ALSKP) and Project MinE ALS sequencing consortium. Secondary structure types were binned based on the following: β-strand/-sheet includes β-strands and β-sheets; helices includes 3_10_-helices, α-helices, π-helices, and polyproline II helices; and coils includes loops, bends, and turns. **(B)** Enrichment analyses were performed using Fishers Exact testing to compare the number of variants carried by individuals with ALS and controls at residues of each secondary structure type in the ALSKP and Project MinE datasets, followed by a Cochran–Mantel–Haenszel (CMH) test. Significance was measured at an alpha-level of 0.05. **(C)** Quantile-quantile plots of rare variants of interest identified across 24 ALS associated genes in ALS case-control sequencing datasets, defined by their secondary structure type. The secondary structure types significantly enriched for variants of interest in the individuals with ALS compared to the controls were further analyzed to determine which genes were driving the enrichment of the features using Fisher’s exact testing. Enrichment of variants in β-strands/-sheets was driven by *SOD1* (ALSKP, p = 6.26e-13; Project MinE, p = 8.38e-03; combined analysis, p = 6.97e-14) and *ANG* (ALSKP, p = 2.04e-03; combined analysis, p = 1.05e-03). Enrichment of variants in helices was driven by *NEK1* (ALSKP, p = 2.92e-03; Project MinE, p = 5.43e-03; combined analysis, p = 4.81e-05), *SQSTM1* (ALSKP, p = 4.92e-05; combined analysis, p = 6.41e-04), *OPTN* (ALSKP, p = 1.65e-03; Project MinE, p = 5.43e-03; combined analysis, p = 9.52e-04). An alpha-level of 2.08e-03 was considered significant following Bonferroni correction accounting for the 24 genes analyzed.

We next assessed the RSA of the reference amino acids involved in the rare, missense variants. Mean RSAs of the variants in any of the 24 genes carried by individuals with ALS and controls in the ALS Knowledge Portal and Project MinE ALS sequencing consortium were compared using Wilcoxon rank sum tests with continuity correction, but no significant differences were observed (ALS Knowledge Portal p-value = 0.142 and Project MinE ALS p-value = 0.1583; Figure 4A). Comparison of mean RSAs were also performed, and only *SOD1* displayed a significant difference in both datasets with a lower mean RSAs in the variants carried by individuals with ALS compared to controls (ALS Knowledge Portal p = 3.17e-09; Project MinE ALS p = 3.03e-02; Figure 4B).

**Figure 4.**
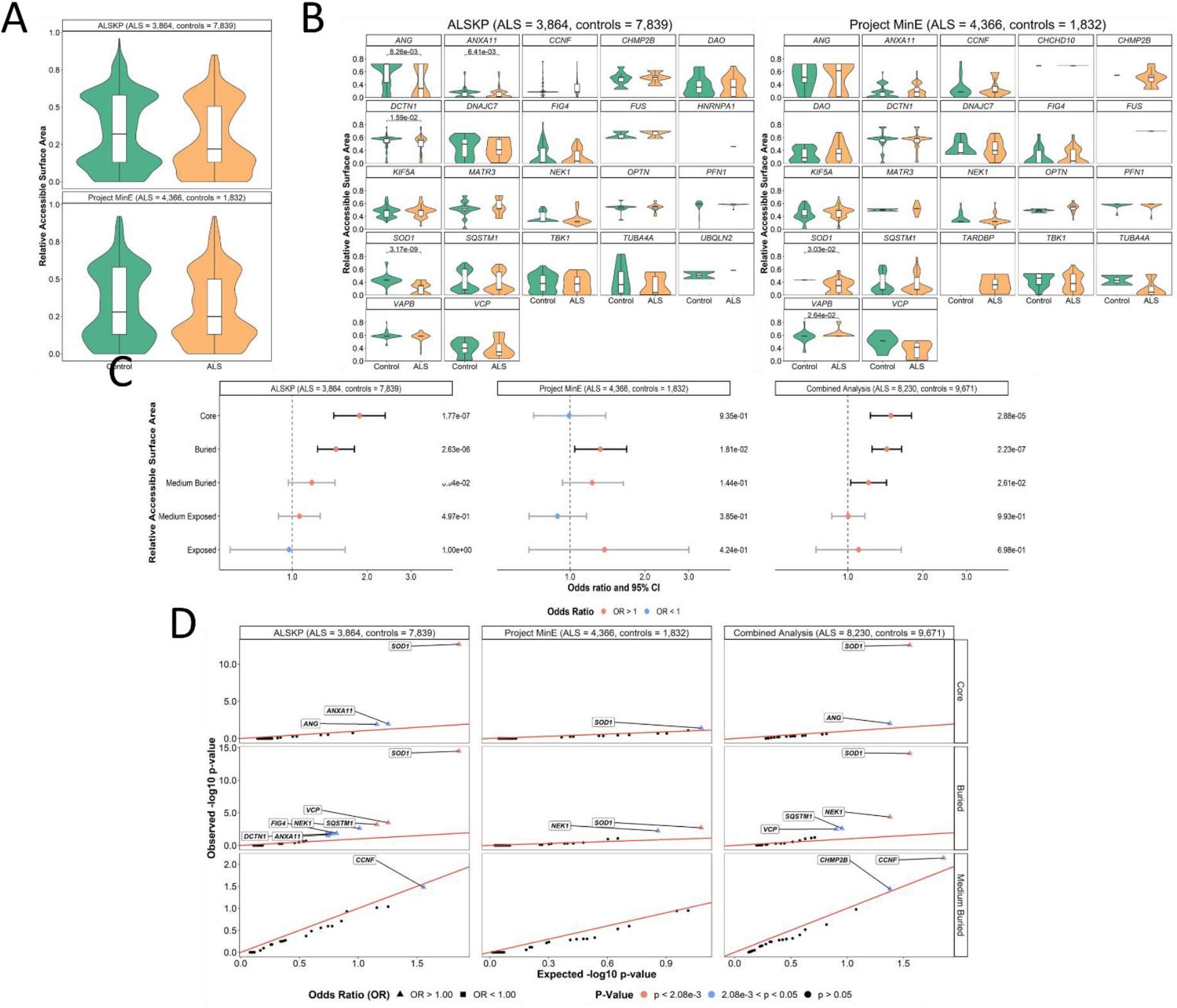
Relative solvent accessible areas (RSAs) of rare missense variants identified across 24 ALS associated genes. For all residues at which variants of interest were observed in the datasets, the AlphaFold predicted structures and DSSP (dictionary of protein secondary structure) program were used to derive RSAs. **(A)** A Wilcoxon rank sum test with continuity correction was used to compare the mean RSA of variants carried by the individuals with ALS and controls from the ALS Knowledge Portal (ALSKP) and Project MinE ALS sequencing consortium. Significant differences were not observed in either dataset (p-value = 0.142 and p-value = 0.1583, respectively). **(B)** Wilcoxon rank sum tests were used to compare the mean RSA of variants carried by the individuals with ALS and controls per gene. Significant p-values are displayed. Genes with < 2 variant counts were excluded. **(C)** Enrichment analyses were performed using Fishers Exact testing to compare the number of variants carried by individuals with ALS and controls of various RSA levels in the two datasets, followed by a Cochran– Mantel–Haenszel (CMH) test. Significance was measured at an alpha-level of 0.05. **(D)** Quantile-quantile plots of rare missense variants identified across 24 ALS associated genes in the two datasets, defined by their RSA level. The RSA levels significantly enriched for missense variants in the individuals with ALS were further analyzed to determine which genes were driving the enrichment of the features using Fisher’s exact testing. Enrichment of variants in the core was driven by *SOD1* (ALSKP, p = 2.11e-13; Project MinE, p = 3.96e-02; combined analysis, p = 2.72e-13), and *ANG* (ALSKP, p = 1.23e-02; combined analysis, p = 9.73e-03). Enrichment of variants in buried regions was driven by *SOD1* (ALSKP, p = 3.38e-15; Project MinE, p = 1.97e-03; combined analysis, p = 7.71e-15), *NEK1* (ALSKP, p = 2.41e-03; combined analysis, p = 4.39e-05), *SQSTM1* (ALSKP, p = 6.13e-04; Project MinE, p = 1.97e-03 ; combined analysis, p = 1.69e-05), and *VCP* (ALSKP, p = 3.26e-04; combined analysis, p = 3.12e-03). An alpha-level of 2.08e-03 was considered significant following Bonferroni correction accounting for the 24 genes analyzed. Solvent area levels were defined as: core, RSA < 5%; buried, 5% < RSA < 25%; medium-buried, 25% < RSA < 50%; medium-exposed, 50% < RSA < 75%; and exposed, RSA > 75%.

RSAs were then binned into five exposure levels, including: core, RSA <5%; buried, 5% < RSA < 25%; medium-buried, 25% < RSA < 50%; medium-exposed, 50% < RSA < 75%; and exposed, RSA > 75%, to determine whether there was an enrichment of specific exposure levels. In the combined dataset analysis, variants across all 24 genes were enriched in individuals with ALS at core (OR = 1.49 [1.23-1.80], p = 2.88e-05), buried state (OR = 1.43 [1.25-1.64], p = 2.23e-07), and medium-buried state (OR = 1.21 [1.03-1.43], p = 2.61e-02) (Figure 4C).

Gene-based analysis revealed that ALS variant enrichment in residues at protein core was driven by *SOD1* (OR = 68.29 [8.70-535.85], p = 2.72e-13); and that at buried level was also driven by *SOD1* (OR = 119.41 [7.37-1935.76], p = 7.71e-15) as well as *NEK1* (OR = 1.93 [1.41-2.63], p = 4.39e-05). No individual genes were significantly enriched for variants of residues at medium-buried residues.

### Variant protein sequence feature assessment

The variants were also annotated with UniProt-defined functional features of the reference amino acids (Figure 5A). We observed a significant enrichment of variants in ALS in the combined dataset analysis at amino acids involved in compositional bias (OR = 3.22 [2.21-4.70], p = 7.79e-10); domains (OR = 1.44 [1.14-1.81], p = 2.47e-03); regions of interest (OR = 1.18 [1.03-1.35], p = 1.68e-02); and zinc fingers (OR = 1.84 [1.04-3.27], p = 4.66e-02; Figure 5B). The specific genes driving the signal in compositionally biased regions were *TARDBP* (OR = 27.66 [5.77-132.52], p = 1.31e-08) and *FUS* (OR = 2.93 [1.74-4.94], p = 6.28e-05); and in regions of interest, was also *TARDBP* (OR = 28.68 [6.02-136.63], p = 5.29e-09) (Figure 5C).

**Figure 5.**
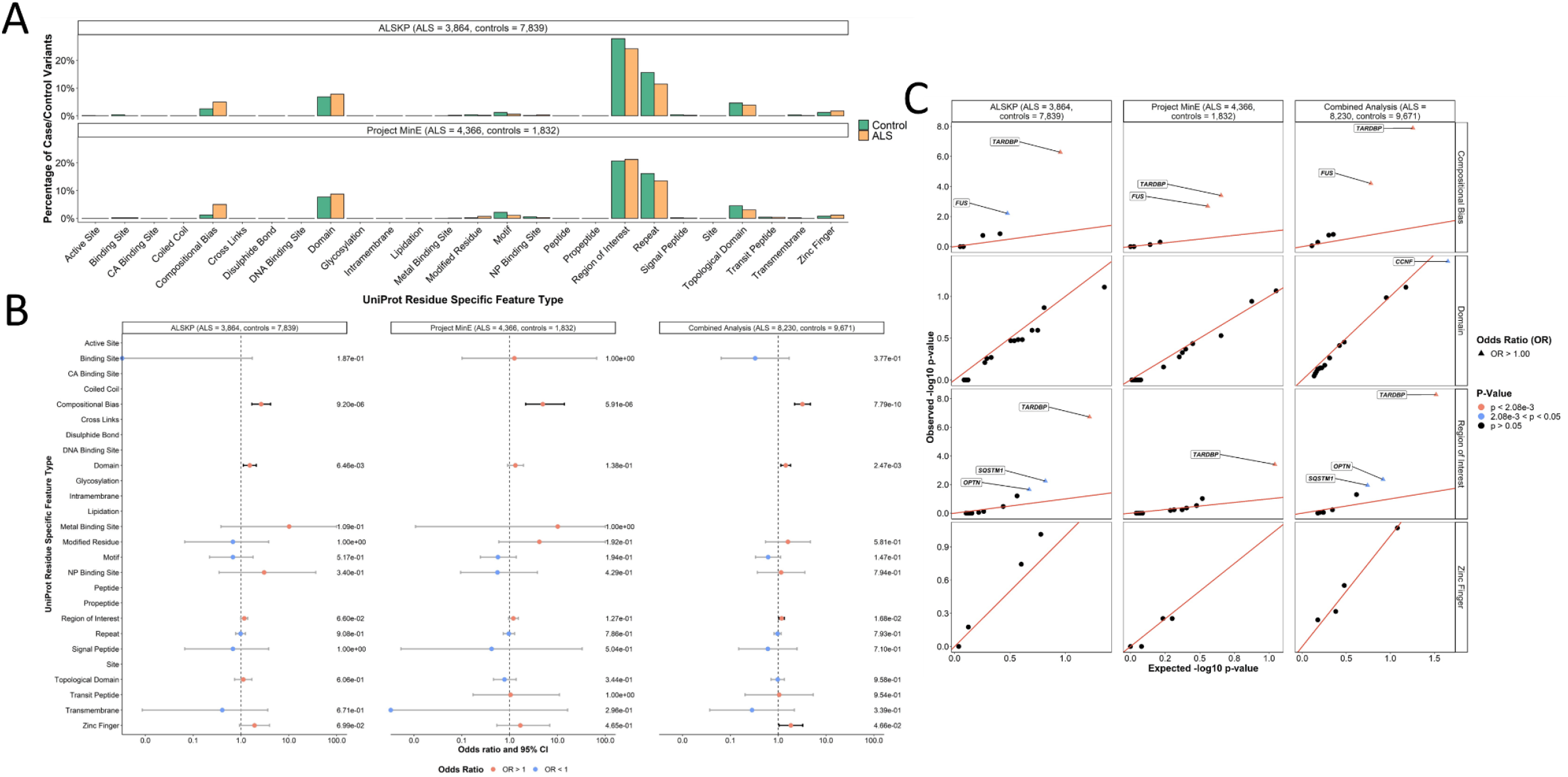
UniProt protein sequence functional features of rare missense variants identified across 24 ALS associated genes. **(A)** Distribution of UniProt defined protein sequence functional features of the residues at which variants of interest were identified carried by the individuals with ALS and controls from the ALS Knowledge Portal (ALSKP) and Project MinE ALS sequencing consortium. **(B)** Enrichment analyses were performed using Fishers Exact testing to compare the number of variants carried by individuals with ALS and controls of UniProt functional features in the ALSKP and Project MinE datasets, followed by a Cochran– Mantel–Haenszel (CMH) test. Significance was measured at an alpha-level of 0.05. **(C)** Quantile-quantile plots of rare variants of interest identified across 24 ALS associated genes in the two datasets, defined by their UniProt functional features. The UniProt functional features significantly enriched for variants of interest in the individuals with ALS compared to the controls were further analyzed to determine which genes were driving the enrichment of the features using Fisher’s exact testing. Enrichment of variants in compositionally biased regions was driven by *TARDBP* (ALSKP, p = 5.23e-07; Project MinE, p = 3.94e-04 ; combined analysis, p = 1.31e-08) and *FUS* (ALSKP, p = 2.03e-03; Project MinE, p = 6.19e-03; combined analysis, p = 6.28e-05). Enrichment of variants in regions of interest was driven by *TARDBP* (ALSKP, p = 1.91e-07; Project MinE, p = 3.94e-04 ; combined analysis, p = 5.29e-09). An alpha-level of 2.08e-03 was considered significant following Bonferroni correction accounting for the 24 genes analyzed.

Similarly, we surveyed the PTM features of the reference amino acids (Figure 6A). We observed a significant enrichment of variants in ALS in the combined dataset analysis at amino acids involved in any known PTM (OR = 1.42 [1.22-1.64], p = 4.87e-06; Figure 6B). This was primarily driven by *SOD1* (OR = 3.57 [2.21-5.76], p = 5.77e-08); *SQSTM1* (OR = 1.65 [1.26-2.16], p = 2.61e-04); *FUS* (OR = 23.19 [2.49-216.23], p = 5.12e-04); and *VCP* (OR = 20.00 [1.15-346.50], p = 7.13e-04) (Figure 6C).

**Figure 6.**
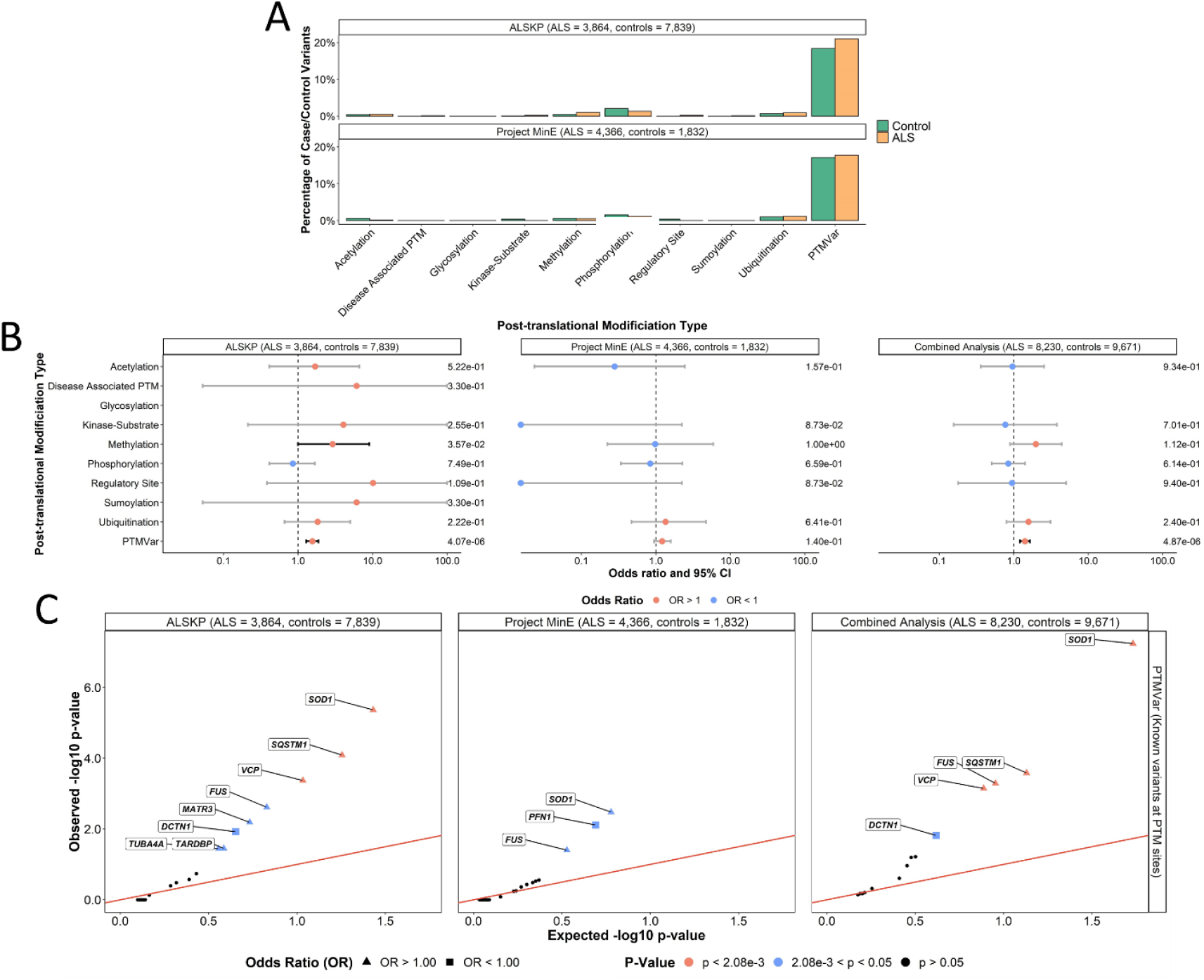
Post-translational modification (PTM) features rare missense variants identified across 24 ALS associated genes. **(A)** Distribution of PTM features of the residues at which all missense variants were identified. **(B)** Enrichment analyses were performed using Fishers Exact testing to compare the number of variants carried by individuals with ALS and controls at which PTMs occur in the ALS Knowledge Portal (ALSKP) and Project MinE datasets, followed by a Cochran–Mantel–Haenszel (CMH) test. Significance was measured at an alpha-level of 0.05. **(C)** Quantile-quantile plots of rare missense variants identified across 24 ALS associated genes in ALS case-control sequencing datasets, that are located at known post-translational modification sites (PTMVar). Known PTM sites, which were significantly enriched for variants of interest in the individuals with ALS compared to the controls from the two datasets, were further analyzed to identify the genes driving the enrichment using Fisher’s exact testing. Enrichment of variants was driven by *SOD1* (ALSKP, p = 4.32e-06; Project MinE, p = 3.33e-03; combined analysis, p = 5.77e-08), *SQSTM1* (ALSKP, p = 8.13e-05; combined analysis, p = 2.61e-04), *FUS* (ALSKP, p = 2.43e-03; Project MinE, p = 3.96e-02; combined analysis, p = 5.12e-04), and *VCP* (ALSKP, p = 4.26e-04; combined analysis, p = 7.13e-04). An alpha-level of 2.08e-03 was considered significant following Bonferroni correction accounting for the 24 genes analyzed.

### Variant transcriptional expression levels

Finally, all variants were annotated with expression levels according GTEx across all tissues and specifically within brain tissues (Figure 7A). We compared the expression levels of the residues and observed a significant enrichment of variants in ALS at amino acids with high and medium expression levels in all tissues (OR = 1.56 [1.38-1.77], p = 3.79e-13 and OR = 1.17 [1.07-1.28], p = 8.41e-04, respectively); and in brain specific tissues (OR = 1.54 [1.37-1.73], p = 1.65e-13 and OR = 1.15 [1.05-1.27], p = 2.38e-03, respectively; Figure 7B). For the high expression in both ‘all tissues’ and in ‘brain tissues’, *SOD1* was the primary driver (OR = 10.49 [5.58-19.70], p = 9.70e-22 and OR = 10.49 [5.58-19.70], p = 9.70e-22, respectively; Figure 7C). Further, variants with medium expression in all tissues were significantly enriched in *OPTN* (OR = 2.52 [1.52-4.16], p = 2.60e-04) and variants with medium expression both in all tissues and in brain tissues were significantly enriched in *FUS* (OR = 2.37 [1.39-4.03], p = 1.94e-03 and OR = 2.37 [1.39-4.03], p = 1.94e-03, respectively) as well as *TARDBP* (OR = 2.63 [1.42-4.87], p = 1.96e-03 and OR = 2.63 [1.42-4.87], p = 1.96e-03, respectively).

**Figure 7.**
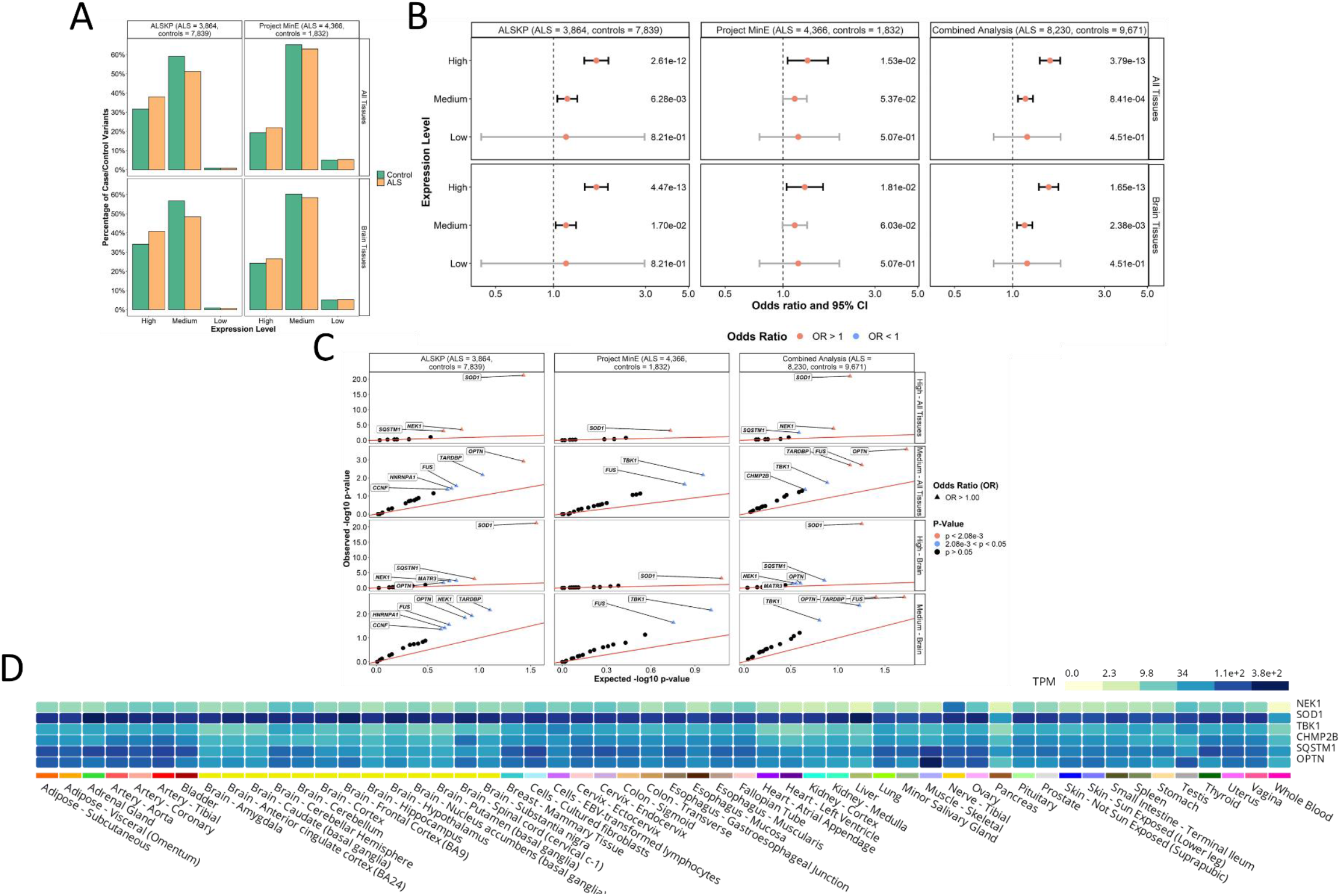
Expression levels of rare missense variants identified across 24 ALS associated genes. **(A)** Distribution of expression levels across all tissues and in brain tissues of all missense variants found in the ALS Knowledge Portal (ALSKP) and Project MinE sequencing consortium datasets were identified. **(B)** Enrichment analyses were performed using Fishers Exact testing to compare the expression levels across all tissues and in brain tissues of variants carried by individuals with ALS and controls of affecting different amino acids in the two datasets, followed by a Cochran–Mantel–Haenszel (CMH) test. Expression levels were defined as: low, mean expression < 0.1; medium, mean expression ≥ 0.1 and ≤ 0.9; and high, mean expression > 0.9. Significance was measured at an alpha-level of 0.05. **(C)** Quantile-quantile plots of missense variants identified across the 24 ALS associated genes, defined by their expression level in brain tissues and all tissues. The expression levels significantly enriched for variants of interest in the individuals with ALS compared to the controls from the ALSKP and Project MinE ALS datasets were further analyzed to determine which genes were driving the enrichment of the features using Fisher’s exact testing. Enrichment of variants with high expression in all tissues was driven by *SOD1* (ALSKP, p = 5.64e-22; Project MinE, p = 6.60e-04; combined analysis, p = 9.70e-22), *NEK1* (ALSKP, p = 3.22e-04; combined analysis, p = 1.52e-04), and *SQSTM1* (ALSKP, p = 1.04e-03; combined analysis, p = 3.30e-03), and enrichment of variants with medium expression in all tissues was driven by *OPTN* (ALSKP, p = 1.22e-03; combined analysis, p = 2.60e-04), *FUS* (ALSKP, p = 2.79e-02; Project MinE, p = 2.27e-02; combined analysis, p = 1.94e-03), and *TARDBP* (ALSKP, p = 6.80e-03; combined analysis, p = 1.96e-03). Similarly, enrichment of variants with high expression in brain tissues was driven by *SOD1* (ALSKP, p = 5.64e-22; Project MinE, p = 6.60e-04; combined analysis, p = 9.70e-22), and enrichment of variants with medium expression in brain tissues was driven by *FUS* (ALSKP, p = 2.79e-02; Project MinE, p = 2.27e-02; combined analysis, p = 1.94e-03), *TARDBP* (ALSKP, p = 6.80e-03; combined analysis, p = 1.96e-03), and *OPTN* (ALSKP, p = 1.44e-02; combined analysis, p = 4.36e-03). An alpha-level of 2.08e-03 was considered significant following Bonferroni correction accounting for the 24 genes analyzed. **(D)** Average expression of the genes identified as driving enrichment of variants with medium and high expression in the brain and all tissues in individuals with ALS compared to controls based on data from the Genotype-Tissue Expression (GTEx) Portal.

### Case Study: SOD1

To highlight the strength of our approach, we present a case study visualizing our results regarding one of the most well-established ALS-associated genes, *SOD1* (Figure 8). All missense variants identified within *SOD1* in individuals with ALS were annotated with ClinVar pathogenicity classifications and the variant positions were mapped onto the AlphaFold predicted protein structure in the G2P Portal. Protein structure, protein sequence, and transcriptomic features were also mapped onto the SOD1 3D protein structure, including the significantly enriched features: β-sheets/-strands, core and buried regions, known sites of PTMs, and high expression in all tissues and in brain tissues.

**Figure 8.**
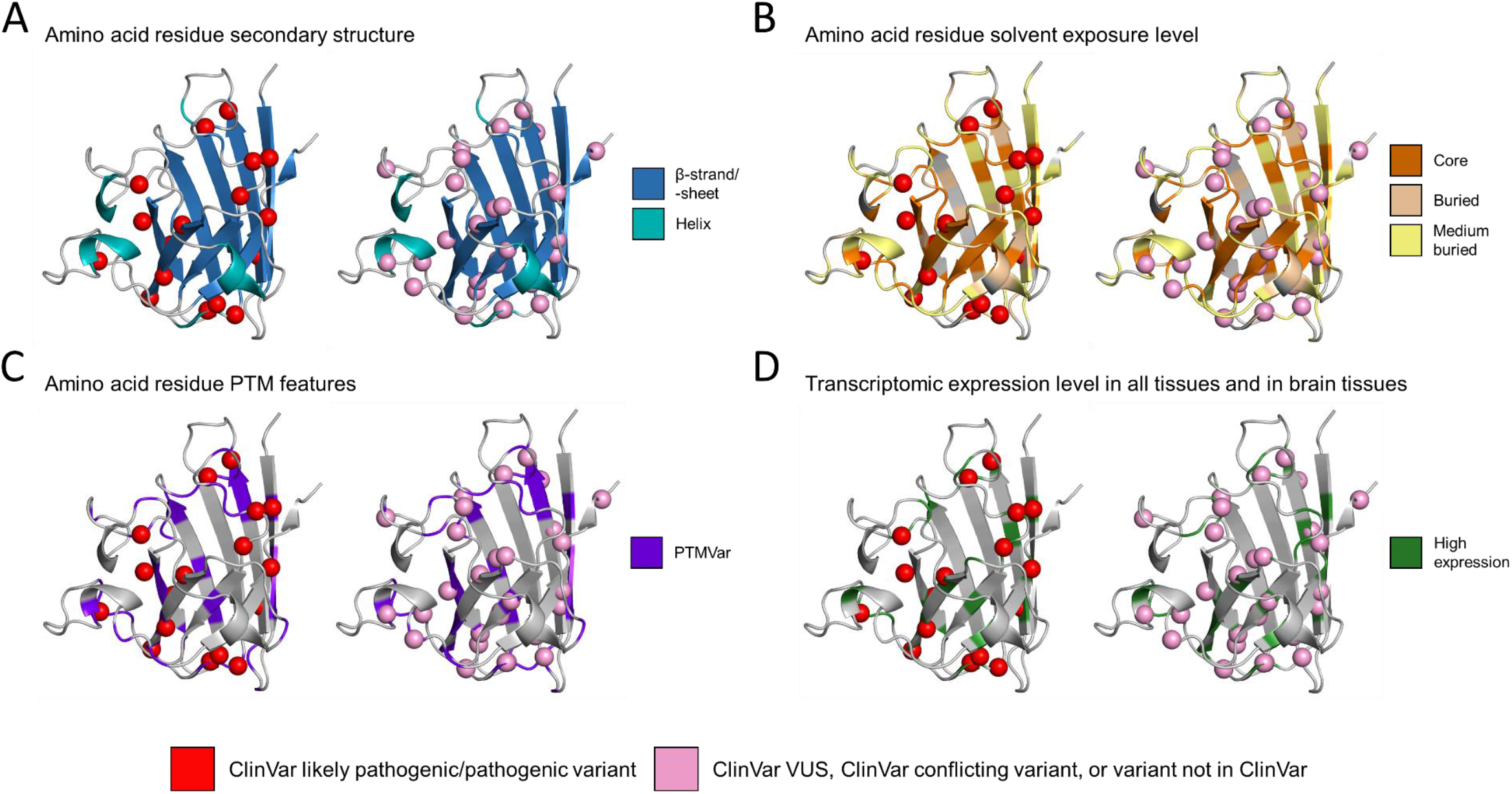
Mapping of missense variants carried by individuals with ALS onto the AlphaFold-predicted structure of SOD1. The Genomics2Protein portal was used to map missense variants carried by the individuals with ALS from the ALS Knowledge Portal and Project MinE ALS sequencing consortium onto the predicted monomeric 3D structures of *SOD1* collected from the AlphaFold database. Missense variants were annotated using the ClinVar database to determine whether they had been previously classified based on their pathogenicity. For the purposes of visualization, all variants classified as a variant of uncertain significance (VUS) or conflicting interpretation by ClinVar or not classified by ClinVar were binned. Following the application of a burden analysis approach to determine whether individual genes were driving protein structure, protein sequence, or transcriptomic feature enrichment, we identified the following features were significantly enriched in *SOD1* missense variation carried by individuals with ALS compared to controls: **(A)** β-strand/-sheets secondary structures; **(B)** core and buried solvent exposure levels; **(C)** known post-translational modification sites (PTM); and **(D)** high transcriptomic expression levels in all tissues and in brain tissues. Abbreviations: PTMVar, any known post-translational modification sites.

Of the 42 unique missense variants identified in *SOD1* in individuals with ALS, 16 (38.1%) were classified as pathogenic or likely pathogenic in ClinVar, all of which were located within at least one of the protein structure, protein sequence, or transcriptomic features found to be enriched in *SOD1* missense variants in the individuals with ALS. Further, 10 (23.8%) of these variants were within a β-strand/-sheet (Figure 8A), 13 (31.0%) were within a core or buried region (Figure 8B), 11 (26.2%) were located at known PTM sites (Figure 8C), all 16 exhibited high expression levels (Figure 8D), and six (14.3%) were located at a position annotated with all four enriched features. Twenty-six (61.9%) of the unique missense variants in *SOD1* carried by individuals with ALS were classified as either a VUS in ClinVar, as having conflicting interpretations in ClinVar, or were not reported in ClinVar. Of these variants with unknown pathogenicity, 23 (54.8%) were located within at least one of the enriched features, and seven (16.7%) were located at a position annotated as all four enriched features.

Interestingly, two missense variants were identified in *SOD1* exclusively in control samples, namely p.Thr136Ile and p.Asp110Tyr. Although both variants were considered to have high expression, neither variant were located within an enriched secondary structure type, an enriched solvent exposure level, nor a known PTM site.

## Discussion

Despite the rapid advancement in gene discovery in ALS, our understanding of the pathogenicity of the variants identified within those genes remains challenging, particularly when faced with missense variation. Here, we have used large-scale sequencing and a case-control enrichment analysis approach to identify patterns of protein structure, protein sequence, and transcriptomic features of missense variants in 24 known ALS-associated genes. Specifically, we observed a significant enrichment of missense variants affecting the positively charged amino acids (Arg, Lys), hydrophobic amino acids (Ile, Leu, Val), and amino acids with special properties (Gly; the smallest amino acid, and Pro; the most rigid amino acid with no free hydrogen) in individuals with ALS compared to controls. Further, using the predicted human protein structures from AlphaFold, we determined that missense variants carried by individuals with ALS were significantly enriched in β-sheets and α-helices, as well as in regions considered to be in the protein’s core, buried, or moderately buried regions. We also identified UniProt-defined protein sequence features significantly enriched in variants in individuals with ALS, such as compositionally biased regions (regions composed of a distinct subset of amino acids) and regions of interest and determined that missense variants at known PTM sites should be considered during variant classification. Assessment of the missense variant expression level based on transcriptomics revealed significant enrichment of variants of high and medium expression across all tissues and within the brain. Together, these results suggest that proteomic and transcriptomic features may improve missense variant pathogenicity classification in ALS genetic testing.

While it could be suggested that the features we found enriched in individuals with ALS would be relevant to pathogenic missense variation across a range of diseases, we hypothesize that our results are specific to ALS. We observed a strong, significant signal of enrichment for missense variants in compositionally biases protein regions (e.g., Gly-rich region in FUS, Gln/Gly/Ser/Tyr-rich and Gly-rich regions in TARDBP; Figure 5), in contrast, a recent assessment of UniProt functional features enriched in 189 neurodevelopmental disease (NDD)-associated genes across >360,000 samples found a depletion of NDD variants in compositionally biased protein regions compared to controls (29). Similarly, protein “regions of interest” that mediates protein-protein interactions or another biological process were significantly enriched in the missense variation carried by individuals with ALS herein, but not in the variation carried by individuals with NDD. One contributing factor to the differences in features between NDD missense variation and ALS variation may be that NDD pathogenic variants often display loss of function properties (46-48), while ALS variants often display gain of function properties (21, 49-51). But additional investigations will be required to determine whether the pathological mechanisms translate to the protein features involved in disease-associated missense variation.

Furthermore, it has been previously shown that functional sites such as active sites, metal, nucleotide-phosphate, co-factor binding sites are enriched for missense variations across 1,330 Mendelian disease-associated genes (41), while we observed no significant association between these functional features and ALS missense variants (Figure 5). These differences between the characteristic protein functional features of neurodevelopmental disorder and Mendelian disease mutations, compared to those we identified for ALS suggest that our results are not generalizable to other Mendelian phenotypes.

Upon identification of features enriched for ALS case variants in all 24 genes, we applied a burden analysis approach to determine whether individual genes were driving feature enrichment. The burden signals observed in *SOD1* were of particular interest, as further demonstrated by the presented case study (Figure 8). Notably, *SOD1* missense variants in individuals with ALS were significantly found to affect hydrophobic amino acid residues Ile and Leu, be in β-sheets/-strands, be in the protein core or buried regions, be in known sites of PTMs, and have high expression in all tissues, as well as within brain specific tissues. Upon further investigation of the identified missense variants in *SOD1* by visualization of the protein structure, we observed that of the total 42 unique missense variants in individuals with ALS, 38.1% were classified as pathogenic or likely pathogenic in the ClinVar database. Further, all pathogenic or likely pathogenic missense variants had at least one feature found to be enriched in *SOD1* missense variants in ALS (Figure 8), even following exclusion of expression levels as all identified *SOD1* variants were considered highly expressed. Functional evidence regarding the pathogenic mechanisms involved in *SOD1*-associated ALS closely complements our identified enriched features. Previous studies have found mutations within core β-sheets are more likely to alter the packing of the protein and result in out-of-register β-sheet oligomers, thought to be important to the cytotoxicity of SOD1 (52, 53). Additionally, mutations impacting the β-sheet structures near edge-strands result in loss of stability and increased susceptibility to aggregation (54). Further evidence suggests variants affecting core and buried residues of SOD1 result in destabilization of interactions imperative for proper folding and in exposure of hydrophobic residues, which again induces aggregation (55-58). Finally, aggregation of SOD1 was found to be prompted due to loss of and alteration in PTMs, such as copper and zinc binding, disulphide formation, phosphorylation, and ubiquitination, among others (59-61). Collectively, the consistency between our findings and ClinVar and functional evidence for *SOD1* present a proof of concept that the identified proteomic and transcriptomic features enriched in ALS may improve the definition of pathogenic variants across all 24 ALS associated genes.

In total, there were 26 unique missense variants identified in individuals with ALS that were considered VUS, conflicting interpretations, or unclassified in the ClinVar database in *SOD1* across the ALS datasets. Of these, 23 were annotated with at least one of the protein sequence and structural features or transcriptomic features significantly enriched in the gene (Figure 8), as outlined above, suggesting these variants may have a greater potential for pathogenicity than one would have previously proposed. In contrast, the *SOD1* variants unique to controls did not display any of the enriched protein sequence or protein structural features. Although we are not recommending that variant pathogenicity classification could be based entirely on any one of the enriched features, we do suspect that incorporating the evidence into pathogenicity classification workflows will improve accuracy, in a similar manner to the incorporation of *in silico* prediction tool-based evidence (11).

In addition to *SOD1*, seven genes displayed significant enrichment of at least one proteomic or transcriptomic feature that could be incorporated into a gene- and disease-specific variant pathogenicity classification structure. This may be especially beneficial for the pathogenicity classification of variants in genes with exceptionally high frequencies of missense VUS, such as *SQSTM1* (Figure 1A, B) (62). Based on our findings, extra consideration should be taken for *SQSTM1* missense variants located within helices, specifically α-helices, or at known PTM sites.

Interestingly, α-helices within the *SQSTM1* encoded protein, p62, are main structural components of two of its primary functional domains, namely the PB1 domain and the UBA ubiquitin-binding domain (63, 64). Further, many post-translational modifications have been observed for p62, including acetylation, disulfide bridging, phosphorylation, and ubiquitination, which are imperative for the protein’s role in autophagy (65-67). As the current ACMG pathogenicity classification guidelines do not account for these gene-specific features, unless the regions are also pre-determined mutational hotspots (11), the incorporation of significant protein structure, protein sequence, and transcriptomic features may improve pathogenicity classification of missense variation found within the genes with large rates of VUS.

There were also significant signals of enrichment that were not driven by specific ALS-associated genes, including for missense variants in moderately buried regions, domains, zinc fingers, or involving a selection of reference amino acids. While the lack of significant results in these burden analyses may have resulted from lack of statistical power due to sample size, the ALS Knowledge Portal and Project MinE ALS sequencing consortium are considered two of the largest ALS sequencing datasets currently available. Further, the number of missense variants that were annotated with these specific features were not exceptionally small; for example, across the two datasets there were a greater number of medium buried missense variants (n = 644) identified than core missense variants (n = 482) for which specific gene burdens were identified. Therefore, it is not unreasonable to propose that features that are not driven by single genes may indeed be relevant across all, or many, ALS-associated genes and warrant further exploration.

There are important caveats to our presented analyses that we acknowledge within our study. The individuals included within the ALS Knowledge Portal and Project MinE ALS sequencing consortium cohorts were largely of European ancestry (30, 31), but it is well established that rates of VUS are much higher in underrepresented populations (68, 69). As larger, more diverse cohorts of ALS patients become available, it will be important to replicate our analyses using those datasets to ensure that our results remain applicable across all populations. The results presented also have yet to be functionally validated using *in vitro* or *in vivo* methodologies; however, as the success of case-control analyses have demonstrated in the identification of novel disease associated genes, the application of large-scale sequencing data is highly opportunistic in the identification of genetic signatures and our study demonstrates how integrating other ‘-omics’ datatypes may expand our understanding of known disease-associated genes. As previously stated, we do not believe that the features we identified can be used as sole evidence for pathogenicity, rather our results suggest that the incorporation of proteomic and transcriptomic evidence may offer additional insights for missense variation.

## Conclusion

In conclusion, our results demonstrate that rare, missense variants identified in two of the largest ALS case-control cohorts to date exhibit enrichment of protein structure, protein sequence, and transcriptomic features that may be useful in defining variant pathogenicity. Although there likely are novel genomic ALS risk factors yet to be uncovered, our findings reveal that some of the missing heritability exhibited by the phenotype may be encompassed by VUS in known ALS-associated genes. Incorporating gene-specific features into variant classification may offer the ability to better identify true pathogenic missense variants in ALS clinical cohorts, thereby improving diagnoses and making clinical trials more accessible to a larger proportion of patients.

## Data Availability

All data produced are available online at

https://ndkp.hugeamp.org/

http://databrowser.projectmine.com/

## Declarations

### Ethics approval

Only summary genetic data from the open-access ALS Knowledge Portal and the open-access Project MinE Databrowser, were used for the analyses described herein. Appropriate and informed consent in accordance with each Research Ethics Board at each respective recruiting site was obtained as described in the primary publications (30), (31).

### Data availability

ALS Knowledge Portal data is available through the Neurodegenerative Disease Knowledge Portal (https://ndkp.hugeamp.org/). Project MinE sequencing consortium data is available through the Project MinE Databrowser (http://databrowser.projectmine.com/). The AlphaFold Protein Structure Database (https://alphafold.ebi.ac.uk/) and Genomics2Protein Resource (https://g2p.broadinstitute.org/) are open access and were used for these analyses.

### Author’s contributions

AAD contributed to the conceptualization, methodology, data curation, formal analyses, visualization, writing of the original draft, and review/editing of the manuscript. GAR contributed to supervision and review/editing of the manuscript. SI contributed to the conceptualization, methodology, data curation, visualization, supervision, writing of the original draft, and review/editing of the manuscript. SMKF contributed to the conceptualization, methodology, supervision, writing of the original draft, and review/editing of the manuscript.

## Acknowledgements

AAD is supported by the Canadian Institute of Health Research Banting Postdoctoral Fellowship program.

## Footnotes

1 http://databrowser.projectmine.com/

2 https://www.uniprot.org/help/sequence_annotation

3 https://g2p.broadinstitute.org/

