## Supplemental for "Characterizing proteomic and transcriptomic features of missense variants in amyotrophic lateral sclerosis genes"

**Supplemental Table 1. Summary of the 24 amyotrophic lateral sclerosis (ALS) associated genes interrogated for rare, missense variation in the exome sequencing datasets from the ALS Knowledge Portal and Project MinE ALS sequencing consortium.**

| **Gene** | **Year of First Association** | **Onset** | **Inheritance Pattern** | **Method of First Association** | **Protein Function** | **PMID of First Association** |
| --- | --- | --- | --- | --- | --- | --- |
| *ANG* | 2006 | Adult | AD | Candidate gene | RNA processing | 16501576 |
| *ANXA11* | 2017 | Adult | AD | Rare variant association (familial/case-control studies) | Intracellular cargo transport | 28469040 |
| *CCNF* | 2016 | Adult | AD | Linkage | Ubiquitination | 27080313 |
| *CHCHD10* | 2014 | Adult | AD | Rare variant association (familial/case-control studies) | Mitochondrial function | 24934289 |
| *CHMP2B* | 2006 | Adult | AD | Candidate gene | Cell maintenance | 16807408 |
| *DAO* | 2010 | Adult | AD | Candidate gene | Stress response | 20368421 |
| *DCTN1* | 2003 | Adult | AD | Candidate gene | Intracellular cargo transport | 12627231 |
| *DNAJC7* | 2019 | Adult | AD | Rare variant association (familial/case-control studies) | Stress response | 31768050 |
| *FIG4* | 2009 | Adult | AD | Candidate gene | Endosomal trafficking | 19118816 |
| *FUS* | 2009 | Adult | AD/AR | Linkage | RNA processing | 19251627 |
| *HNRNPA1* | 2013 | Adult | AD | Rare variant association (familial/case-control studies) | RNA processing | 23455423 |
| *KIF5A* | 2018 | Adult | AD | Rare variant association (familial/case-control studies) | Cytoskeleton | 29342275 |
| *MATR3* | 2014 | Adult | AD | Rare variant association (familial/case-control studies) | RNA processing | 24686783 |
| *NEK1* | 2016 | Adult | AD | Rare variant association (familial/case-control studies) | Cell maintenance | 26945885 |
| *OPTN* | 2010 | Adult | AD | Linkage | Cell maintenance | 20428114 |
| *PFN1* | 2012 | Adult | AD | Rare variant association (familial/case-control studies) | Cytoskeleton | 22801503 |
| *SOD1* | 1993 | Adult | AD | Linkage | Stress response | 8446170 |
| *SQSTM1* | 2011 | Adult | AD | Candidate gene | Cell maintenance | 22084127 |
| *TARDBP* | 2008 | Adult | AD | Linkage | RNA processing | 18309045 |
| *TBK1* | 2015 | Adult | AD | Rare variant association (familial/case-control studies) | Cell maintenance | 25700176 |
| *TUBA4A* | 2014 | Adult | AD | Rare variant association (familial/case-control studies) | Cytoskeleton | 25374348 |
| *UBQLN2* | 2011 | Adult | X-LD | Linkage | Cell maintenance | 21857683 |
| *VAPB* | 2004 | Adult | AD | Linkage | Stress response | 15372378 |
| *VCP* | 2010 | Adult | AD | Rare variant association (familial/case-control studies) | Cell maintenance | 21145000 |

Abbreviations: AD, autosomal dominant; AR, autosomal recessive; PMID, PubMed ID; X-LD, X-linked dominant.

**Supplemental Table 2. Protein sequence functional feature annotations from the UniProt database used to annotate the amino acid residues substituted in the analysed missense variants.**

|  | **Functional site annotation** | | **Definition** |
| --- | --- | --- | --- |
| Active site | | Amino acid is involved in the activity of an enzyme | |
| Binding site | | Amino acid is involved in the binding site of a chemical group | |
| Calcium binding site | | Amino acid is involved in the binding site of calcium | |
| Coiled coil | | Amino acid is in a region of a coiled coil in the protein | |
| Compositional bias | | Amino acid is in a region of protein made up of mainly a distinct subset of amino acids | |
| Cross link | | Amino acid participates in covalent linkage(s) with another amino acid | |
| Disulphide bond | | Cysteine residue that participates in disulphide bonding with another cysteine residue | |
| DNA binding site | | Amino acid is in a domain that participates in DNA binding | |
| Domain | | Amino acid is in region considered to be a modular protein domain | |
| Glycosylation | | Amino acid participates in covalently attaching glycan group(s) | |
| Intramembrane | | Amino acid is in a region that is in a membrane, but does not cross it fully | |
| Lipidation | | Amino acid participates in covalently attaching lipid group(s) | |
| Metal binding site | | Amino acid is involved in the binding site of metal ion(s) | |
| Modified residue | | Amino acid is involved in a modification excluding lipids, glycans, and protein cross links | |
| Motif | | Amino acid is in a short sequence motif (≤20 amino acids) of biological interest | |
| Nucleoprotein binding site | | Amino acid is involved in the binding site of nucleotprotein(s) | |
| Peptide | | Amino acid is included in the active peptide, which describes a small polypeptide ((≤50 amino acids) processed from a larger precurser protein that has a defined biological activity | |
| Propeptide | | Amino acid is included in the part of the protein cleaved from the active peptide during maturation or activation | |
| Region of interest | | Amino acid is in a region that has been experimentally defined, such as the role of a region in mediating protein-protein interactions or some other biological process | |
| Repeat | | Amino acid is in a region of repeated sequence motifs or repeated domains | |
| Signal peptide | | Amino acid is in a region of sequence that targets the protein to the secretory pathway or periplasmic space | |
| Site | | Amino acid is considered to be at a site of interest for any reason | |
| Topological domain | | Amino acid is in a non-membrane region of a membrane-spanning protein | |
| Transit peptide | | Amino acid is included in a peptide responsible for transport of a protein to a particular organelle | |
| Transmembrane | | Amino acid is in a region that spans an entire membrane | |
| Zinc finger | | Amino acid is in a region involved in a zinc finger conformation | |

Modified from <https://www.uniprot.org/help/sequence_annotation>.

**Supplemental Table 3. Protein sequence post-translational modification (PTM) feature annotations from the PhosphoSitePlus database used to annotate the amino acid residues substituted in the analysed missense variants.**

| **PTM site annotation** | **Definition** |
| --- | --- |
| Acetylation | Amino acid is modified by the attachment of an acetyl group |
| Methylation | Amino acid is modified by the attachment of a methyl group |
| Phosphorylation | Amino acid is modified by the attachment of a phosphate moiety |
| Ubiquitination | Lysine residue is modified by the attachment of ubiquitin |
| Sumoylation | Lysine residue is modified by the attachment of a small ubiquitin-like modifier (SUMO) |
| Glycosylation | Amino acid is modified by the attachment of a carbohydrate moiety |
| Regulatory site | PTM sites that regulate molecular functions, biological processes, and molecular interactions including protein-protein interactions |
| Kinase-substrate | Amino acid is a substrate phosphorylated by a kinase |
| Disease-associated PTM | PTM sites shown to correlate with specific disease states |
| PTMVar | PTM sites (phosphorlyation, ubiquitylation, acetylation, methylation and succinylation) that overlap with genetic variants associated with diseases and genetic polymorphisms. |

Modified from <https://www.phosphosite.org/homeAction>. Abbreviations: PTM, post-translational modification; SUMO, small ubiquitin-like modifier.

**Supplemental Table 4. Gene-wise case-control counts of variants identified across 24 amyotrophic lateral sclerosis (ALS) associated genes in the ALS Knowledge Portal (ALSKP) and Project MinE ALS sequencing consortium (ProjMinE) datasets.**

| **Gene** | **ALSKP Case** | **ALSKP Control** | **ProjMinE Case** | **ProjMinE Control** |
| --- | --- | --- | --- | --- |
| *ANG* | 31 | 62 | 27 | 6 |
| *ANXA11* | 117 | 226 | 283 | 111 |
| *CCNF* | 76 | 114 | 59 | 26 |
| *CHCHD10* | 0 | 0 | 50 | 22 |
| *CHMP2B* | 11 | 13 | 17 | 1 |
| *DAO* | 21 | 55 | 34 | 10 |
| *DCTN1* | 107 | 245 | 180 | 86 |
| *DNAJC7* | 6 | 24 | 37 | 11 |
| *FIG4* | 46 | 69 | 52 | 19 |
| *FUS* | 25 | 21 | 37 | 4 |
| *HNRNPA1* | 9 | 6 | 8 | 2 |
| *KIF5A* | 65 | 121 | 75 | 28 |
| *MATR3* | 36 | 48 | 40 | 14 |
| *NEK1* | 78 | 94 | 112 | 35 |
| *OPTN* | 28 | 26 | 36 | 8 |
| *PFN1* | 23 | 34 | 15 | 17 |
| *SOD1* | 77 | 28 | 51 | 4 |
| *SQSTM1* | 116 | 153 | 133 | 52 |
| *TARDBP* | 19 | 9 | 34 | 6 |
| *TBK1* | 23 | 38 | 61 | 15 |
| *TUBA4A* | 8 | 8 | 4 | 2 |
| *UBQLN2* | 9 | 13 | 0 | 0 |
| *VAPB* | 39 | 74 | 43 | 24 |
| *VCP* | 16 | 19 | 14 | 5 |

Variants were filtered to only include missense variants that were rare in the general population (allele frequency < 0.01 in the gnomAD v2.2.2 NFE non-neurological cohort).

**Supplemental Table 5. Quality assessment of the Alpha-Fold predicted structures of 24 amyotrophic lateral sclerosis (ALS) associated genes.**

| **Gene** | **uniprotID** | **Protein length** | **Very low** | **Low** | **Confident** | **High confident** |
| --- | --- | --- | --- | --- | --- | --- |
| *ANG* | P03950 | 147 | 1% | 17% | 2% | 80% |
| *ANXA11* | P50995 | 505 | 33% | 4% | 3% | 60% |
| *CCNF* | P41002 | 786 | 33% | 3% | 15% | 49% |
| *CHMP2B* | Q9UQN3 | 213 | 15% | 15% | 31% | 38% |
| *CHCHD10* | Q8WYQ3 | 142 | 31% | 42% | 18% | 9% |
| *DAO* | P14920 | 347 | 0% | 1% | 5% | 93% |
| *DCTN1* | Q14203 | 1278 | 16% | 6% | 50% | 28% |
| *DNAJC7* | Q99615 | 494 | 7% | 3% | 9% | 81% |
| *FIG4* | Q92562 | 907 | 13% | 10% | 27% | 50% |
| *FUS* | P35637 | 526 | 69% | 7% | 15% | 8% |
| *HNRNPA1* | P09651 | 372 | 47% | 5% | 9% | 40% |
| *KIF5A* | Q12840 | 1032 | 18% | 6% | 47% | 29% |
| *MATR3* | P43243 | 847 | 53% | 10% | 35% | 1% |
| *NEK1* | Q96PY6 | 1258 | 49% | 7% | 28% | 16% |
| *OPTN* | Q96CV9 | 577 | 23% | 7% | 19% | 52% |
| *PFN1* | P07737 | 140 | 1% | 1% | 6% | 93% |
| *SOD1* | P00441 | 154 | 0% | 0% | 3% | 97% |
| *SQSTM1* | Q13501 | 440 | 37% | 14% | 19% | 30% |
| *TARDBP* | Q13148 | 414 | 40% | 8% | 49% | 4% |
| *TBK1* | Q9UHD2 | 729 | 4% | 5% | 20% | 71% |
| *TUBA4A* | P68366 | 448 | 2% | 3% | 13% | 82% |
| *UBQLN2* | Q9UHD9 | 624 | 32% | 36% | 29% | 3% |
| *VAPB* | O95292 | 243 | 21% | 9% | 31% | 40% |
| *VCP* | P55072 | 806 | 7% | 7% | 43% | 43% |

Values given represent the percentage of the protein length with that level of quality. Quality levels of the Alpha-Fold predicted structures were defined as: very low quality: pLDDT ≤ 50; low quality: 50 < pLDDT ≤ 70; confident in quality: 70 < pLDDT ≤ 90; very high quality: pLDDT > 90.

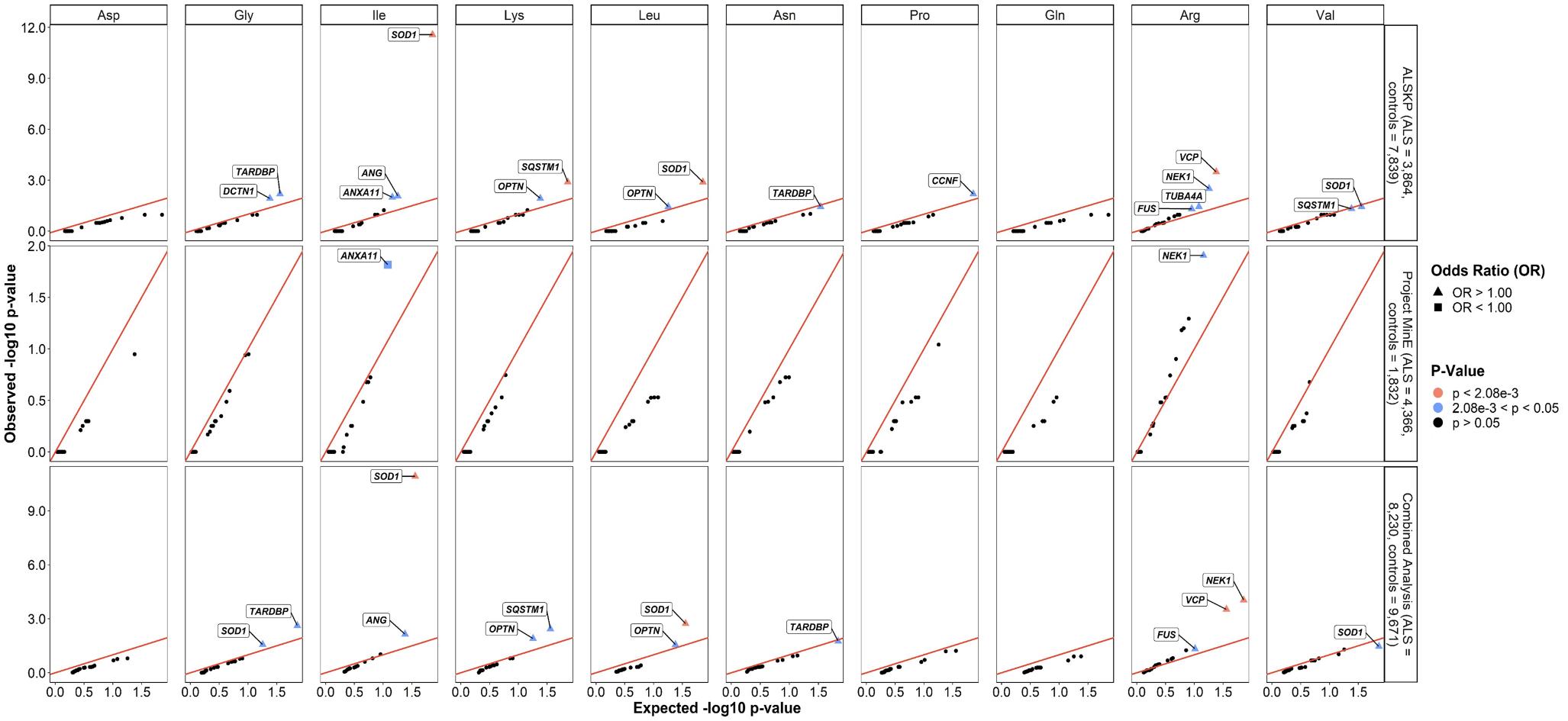
**Supplemental Figure 1. Quantile-quantile plots of rare missense variants identified across 24 ALS associated genes, defined by the reference amino acid.** The reference amino acids significantly enriched for variants of interest in the individuals with ALS compared to the controls from the ALS Knowledge Portal (ALSKP) and Project MinE ALS sequencing consortium datasets were further analyzed to determine which genes were driving the enrichment of the features using Fisher’s exact testing. Enrichment of variants affecting isoleucine residues was driven by *SOD1* (ALSKP, p = 2.68e-12; combined analysis, p = 1.19e-11); enrichment of variants affecting leucine residues was driven by *SOD1* (ALSKP, p = 1.29e-03 ; combined analysis, p = 1.85e-03); enrichment of variants affecting arginine residues was driven by *NEK1* (ALSKP, p = 3.11e-03; Project MinE, p = 1.24e-02; combined analysis, p = 9.15e-05), and *VCP* (ALSKP, p = 3.26e-04; combined analysis, p = 3.01e-04). An alpha-level of 2.08e-03 was considered significant following Bonferroni correction accounting for the 24 genes analyzed.

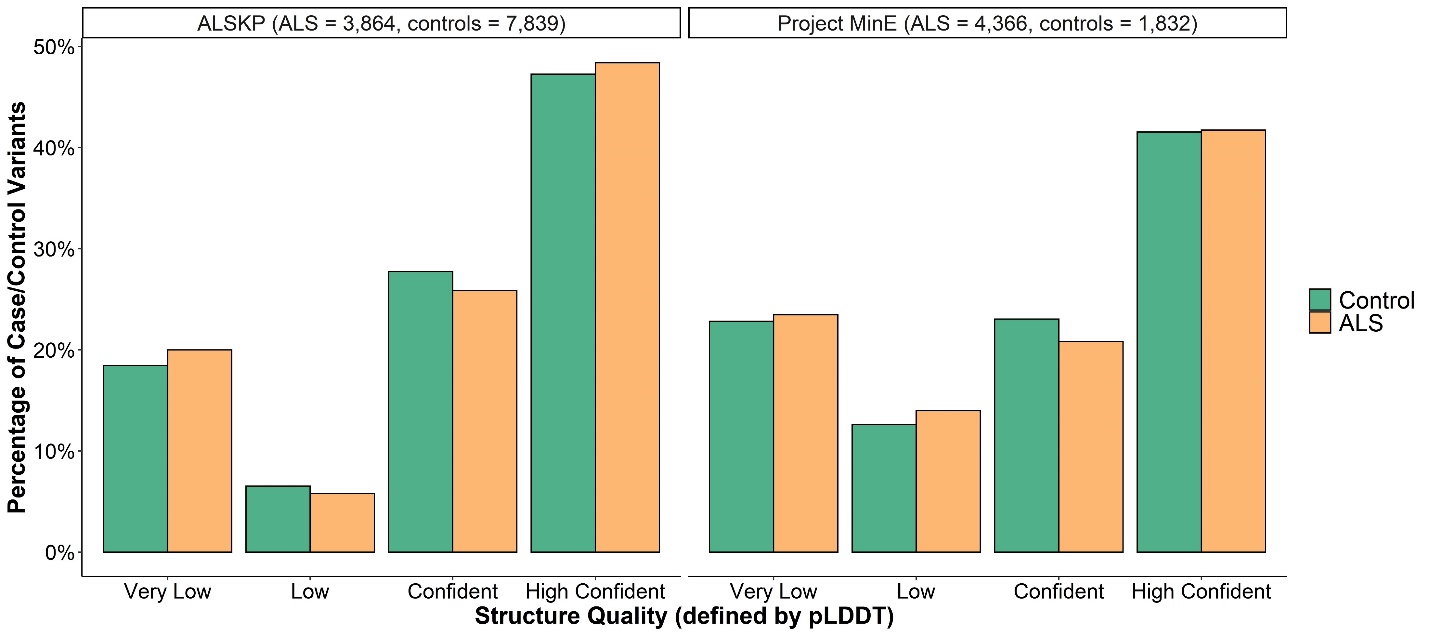

**Supplemental Figure 2. Structure quality of the rare missense variants identified across 24 ALS associated genes in the ALS Knowledge Portal (ALSKP) and Project MinE ALS sequencing consortium datasets.** Values given represent the percentage of variants carried by either an individual with ALS or control at residues with that level of quality. Quality levels of the Alpha-Fold predicted structures were defined using the predicted local-distance difference test (pLDDT), as: very low quality: pLDDT ≤ 50; low quality: 50 < pLDDT ≤ 70; high in quality: 70 < pLDDT ≤ 90; very high quality: pLDDT > 90.

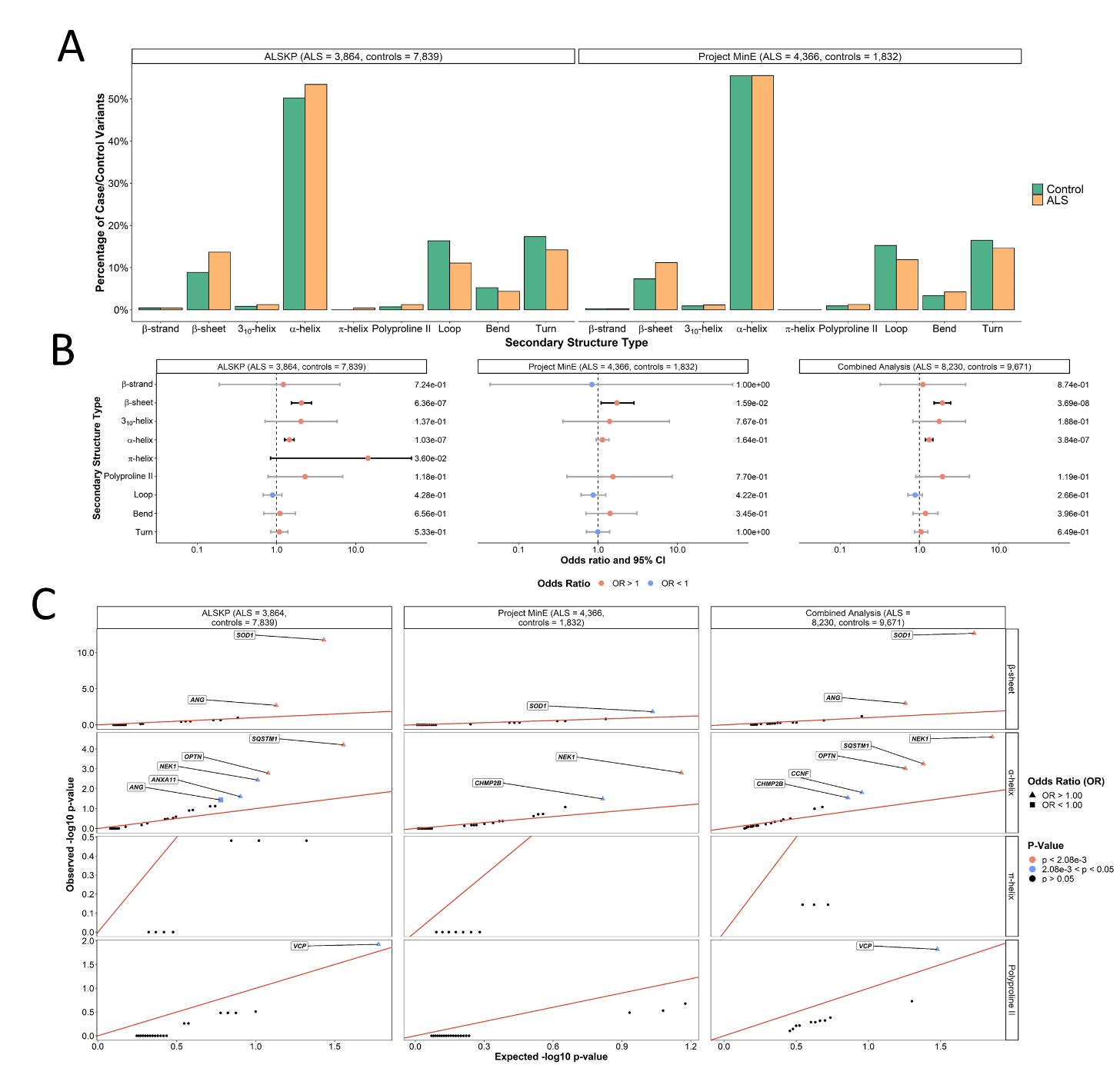

**Supplemental Figure 3. Secondary structure types of the rare missense variants identified across 24 ALS associated genes. (A)** Secondary structure types were obtained from the Alpha-Fold predicted structures for all residues at which variants of interest were observed in the ALS Knowledge Portal (ALSKP) and Project MinE ALS sequencing consortium. **(B)** An enrichment analysis was performed using Fishers Exact testing to compare the number of variants carried by individuals with ALS and controls at residues of each secondary structure type in the ALSKP and Project MinE datasets, followed by a Cochran–Mantel–Haenszel (CMH) test. Significance was measured at an alpha-level of 0.05. **(C)** Quantile-quantile plots of rare variants of interest identified across 24 ALS associated genes in ALS case-control sequencing datasets, defined by their secondary structure type. The secondary structure types significantly enriched for variants of interest in the individuals with ALS compared to the controls from the ALSKP and Project MinE ALS sequencing consortium datasets were further analyzed to determine which genes were driving the enrichment of the features using Fisher’s exact testing. Using an alpha-level of 0.05, enrichment of variants in β-sheets was driven by *SOD1* (ALSKP, p = 1.71e-12; Project MinE, p = 1.47e-02; combined analysis, p = 2.12e-13) and *ANG* (ALSKP, p = 2.04e-03; combined analysis, p = 1.05e-03). Enrichment of variants in α-helices was driven by *NEK1* (ALSKP, p = 3.64e-03; Project MinE, p = 1.58e-03; combined analysis, p = 2.50e-05), *SQSTM1* (ALSKP, p = 6.20e-05; combined analysis, p = 5.61e-04), and *OPTN* (ALSKP, p = 1.65e-03; combined analysis, p = 9.52e-04). An alpha-level of 2.08e-03 was considered significant following Bonferroni correction accounting for the 24 genes analyzed.
